# Risk-benefit assessment of oral anticoagulation in atrial fibrillation using virtual clinical trials

**DOI:** 10.64898/2026.11.04.26367324

**Authors:** Minsi Cai, Fateme Farahani, Michiel Rienstra, Stefano Severi, Harry J.G.M. Crijns, Ulrich Schotten, Jordi Heijman

**Author notes:** Corresponding author: Jordi Heijman, PhD, Gottfried Schatz Research Center, Division of Medical Physics & Biophysics, Medical University of Graz, Neue Stiftingtalstr. 6; 8010 Graz, Austria.

## Abstract

**Background:** Oral anticoagulation (OAC) reduces stroke risk in patients with atrial fibrillation (AF), and screening can enable earlier AF detection and OAC initiation. However, the population-level trade-off between stroke prevention and OAC-related major bleeding for different types of AF and OAC therapy remains uncertain, and large randomized trials to resolve this are costly and time-consuming.

**Methods:** A state-transition Markov-like model was developed to: 1) simulate the lifetime course of AF, stroke, major bleeding, and death in different virtual populations; 2) quantify benefits (stroke reduction) and harms (major bleeding) of systematic AF screening with screening-triggered OAC across CHA_2_DS_2_-VA scores; and 3) compare rhythm-guided, time-delimited (“pill-in-pocket”) OAC with continuous OAC.

**Results:** The model synthesized information from multiple clinical sources and reproduced stroke and bleeding incidence across clinical scenarios. At CHA₂DS₂-VA=2, long-term continuous rhythm monitoring (LT-CRM) with OAC lowered stroke risk (HR, 0.92 [95%CI, 0.88-0.96]) but raised bleeding risk (HR, 1.17 [95%CI, 1.07-1.27]) versus no screening. Time-to-benefit was shorter than time-to-harm for CHA₂DS₂-VA≥4, but longer for CHA₂DS₂-VA=0. Pill-in-pocket OAC vs continuous OAC showed no difference in stroke/mortality (HR, 1.03, 95%CI [0.91-1.16]) but lower bleeding risk (HR, 0.81, 95%CI: 0.68-0.96]) in patients with elevated bleeding risk, but not at baseline. Sensitivity analyses revealed that the risk-benefit profiles of screening or pill-in-pocket OAC strategy further depend on OAC adherence and bleeding risks.

**Conclusions:** Our model highlights a trade-off between stroke reduction and bleeding for systematic screening-initiated OAC, with net benefit for CHA₂DS₂-VA≥4. We provide the first virtual implementation of an ongoing rhythm-guided OAC trial, showing that pill-in-pocket OAC may lower bleeding risk compared to continuous OAC, highlighting the potential of virtual clinical trials to guide clinical trial design.

## Introduction

Atrial fibrillation (AF) is associated with a 2.3-fold increase in ischemic stroke risk.^1^ Oral anticoagulation (OAC) is a cornerstone of AF management, reducing the risk of stroke or thromboembolic events (TE) by 64-80%.^2, 3^ However, the timely diagnosis of AF is challenging due to the highly dynamic and heterogeneous occurrence of AF episodes and weak symptom-rhythm correlation, often precluding timely initiation of OAC. In theory, systematic AF screening would be expected to facilitate early detection of AF, enabling timely OAC initiation and potentially reducing the incidence of stroke/TE. In agreement, several randomized controlled clinical trials (RCTs) have shown increased AF detection and subsequent OAC use with systematic screening.^4–9^ However, to date, no AF-screening studies have demonstrated a specific reduction in stroke/TE,^4, 6, 7^ although one reported a slight reduction in the composite endpoint of ischemic/hemorrhagic stroke, systemic embolism, bleeding leading to hospitalization, and all-cause mortality.^9^ In patients with subclinical AF detected by implantable cardiac devices, OAC was associated with a significantly higher risk of major bleeding.^10–12^ These findings highlight the uncertain risk-benefit profiles of systematic AF screening with subsequent continuous OAC.^13^ Rhythm-guided, time-delimited anticoagulation strategies (“pill-in-pocket” OAC) enabled by recent advances in wearable technologies may offer opportunities to mitigate bleeding risk, but their long-term safety and effectiveness remain unknown and are currently being tested in ongoing RCTs (e.g., Rhythm Evaluation for AntiCoagulaTion with Continuous Monitoring of Atrial Fibrillation [REACT-AF], ClinicalTrials.gov NCT05836987).

While RCTs remain the gold standard to address such therapeutic uncertainties, the high medical costs, long timelines, and ethical constraints of large-scale screening studies make rapid answers based on real-world studies impractical or even impossible. In contrast, computational modeling offers precise control of variables and the ability to simulate complicated disease dynamics, which provides an efficient and ethical alternative for comprehensive intervention assessment.^14–16^ We therefore developed a state-of-the-art computational framework to address the abovementioned unresolved clinical and public health questions using virtual clinical trials. In particular, our study aims to i): evaluate the benefits and harms of systematic AF screening and subsequent OAC; ii) characterize how baseline stroke risk modifies time-to-benefit and time-to-harm of systematic AF screening-initiated OAC; and iii): explore whether a “pill-in-pocket” OAC strategy can improve the risk-benefit profile of OAC following AF detection.

## Methods

### Patient-level model of AF

The current framework builds on our recently developed Markov-like patient-level computational model comprising 7 different clinical states, including pre– or post-stroke sinus rhythm (SR), pre-or post-stroke symptomatic and asymptomatic AF (sAF and aAF), and death.^17^ The model was extended with an explicit representation of the stroke risk, represented by the CHA_2_DS_2_-VA(Sc) scores, and a new parallel component capturing non-bleeding and major bleeding states (**Figure 1A; Figure S1**). OAC initiation decreased the probability of switching to stroke/TE states and increased the probability of switching to the major bleeding state. Entering the major bleeding state caused an immediate and persistent increase of the probability of death. Key assumptions of this model are listed in **Table S1**. Parameter optimization, model calibration, and independent validation are further described in the **Supplemental methods** and **Tables S2-S3**.

**Fig. 1.**
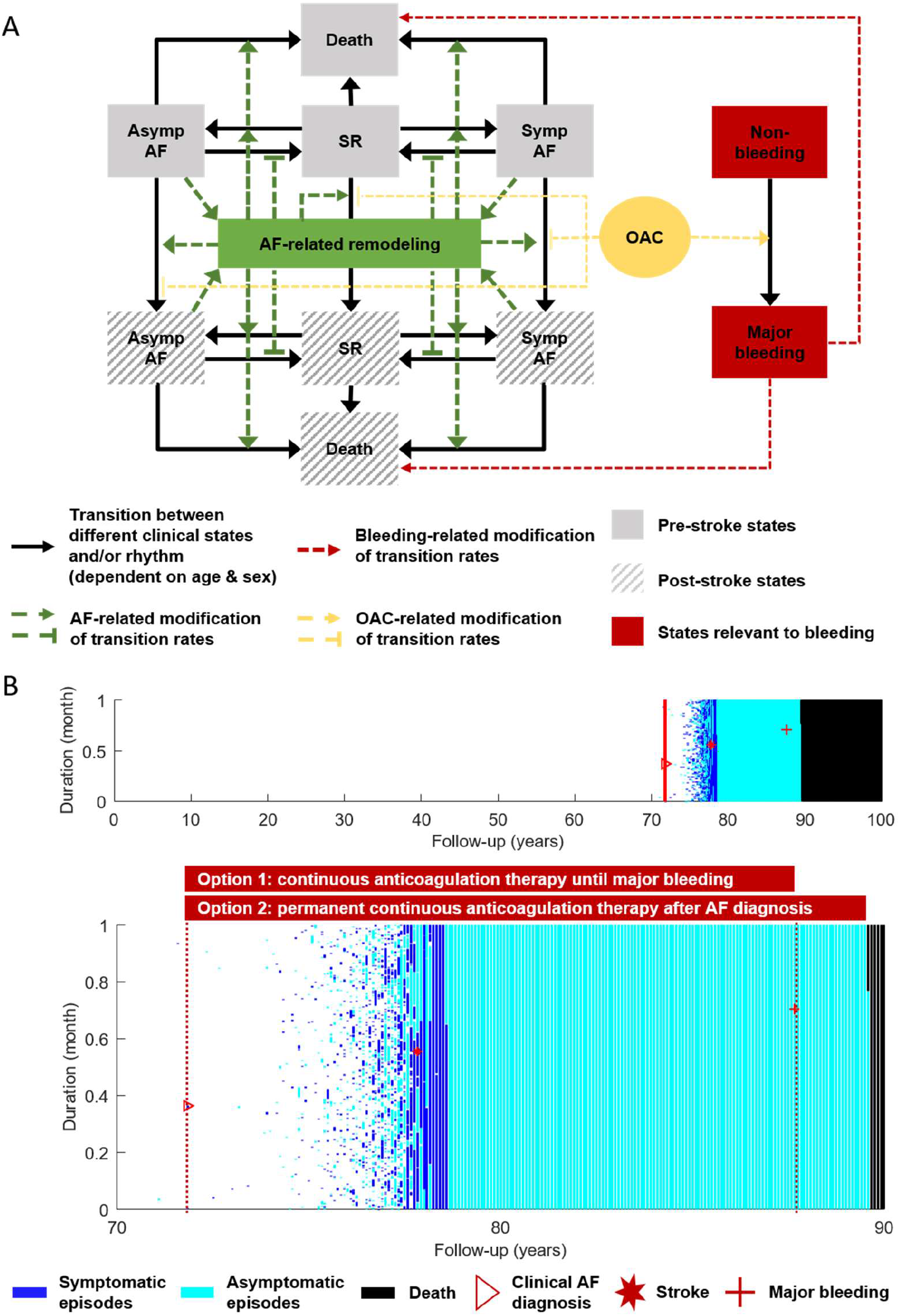
Schematic overview of the Markov-like patient-level model and an example virtual patient with atrial fibrillation (AF) episodes and clinical outcomes. A: Two Markov-like models were integrated, one with 7 clinical states including sinus rhythm, symptomatic AF, and asymptomatic AF, each with or without previous stroke, as well as death. The other one represents non-bleeding and major bleeding states. Transition probabilities were age/sex dependent. AF-related remodeling was also incorporated in the model as function of the time in AF, and modulated the likelihood of AF development, cardioversion, and the occurrence of stroke and death. Oral anticoagulant (OAC) usage was simulated after confirmed clinical AF diagnosis or screening-detected AF by increasing the incidence of major bleeding and reducing the stroke risk. The occurrence of major bleeding would increase the mortality risk. B: An example virtual patient with multiple symptomatic or asymptomatic AF episodes, stroke event, major bleeding event, and death during 100-year follow-up (Top). The bottom panel shows an expanded view from 70 to 90 years of age with progression from paroxysmal AF to persistent and long-standing persistent AF. Continuous OAC was initiated after clinical AF diagnosis (red triangle), and both stroke (red asterisk) and major bleeding (red cross) events occurred afterwards. The length of the bar represented the duration of each AF episode and a complete vertical bar represents 1 month.

### Risk-benefit assessment of AF screening

Three systematic AF screening protocols were simulated: Long-term continuous rhythm monitoring (LT-CRM, e.g., based on implantable loop recorders), yearly 14-day Holter monitoring, and intensive (wearable-based) intermittent screening based on thrice-daily ECGs (**Table S4**). OAC was initiated immediately for virtual patients with either clinical AF (defined as a symptomatic AF episode lasting ≥3 hours) or screening-detected AF. Upon a clinical AF diagnosis, the virtual patient would no longer be eligible for AF screening. Stroke and major bleeding incidence in control and screening groups were compared using Kaplan-Meier (K-M) curves and Cox regression. The main analysis was conducted in a virtual population with an average CHA_2_DS_2_-VA score of 2, and sensitivity analyses were conducted in populations with average CHA_2_DS_2_-VA scores of 0-5, and with real-world OAC adherence (66%), screening sensitivity (95%), and permanent OAC discontinuation after major bleeding. Time-to-benefit (TTB) and time-to-harm (TTH) were calculated at a threshold of 1/500=0.002 (reflecting how many years are needed to observe 1 fewer stroke event and 1 more major bleeding event per 500 individuals screened) to quantify how fast the benefits and harms of AF screening could be incurred, as previously reported.^18, 19^

### Predicting efficacy and safety of pill-in-pocket OAC therapy

The REACT-AF trial (NCT05836987) aims to test the efficacy and safety of intermittent OAC guided by continuous smartwatch-based AF monitoring.^21^ Patients with paroxysmal or persistent AF and CHA_2_DS_2_-VASc scores of 1 to 4 (for males) or 2 to 4 (for females) will be randomized to an intervention group receiving time-delimited OAC guided by an AF-sensing smartwatch or a control group receiving standard continuous OAC. Non-inferiority of the primary outcome (stroke and all-cause mortality) and superiority of the secondary outcome (major-bleeding events) will be tested after 5 years follow-up. The trial is scheduled to be completed in 2029. The comprehensive knowledge of all AF episodes and outcomes during lifetime follow-up for every individual enabled us to implement REACT-AF as a virtual RCT in the current model. The parameter settings and virtual patient selection are provided in **Table S5,** mimicking the protocols of REACT-AF as closely as possible in a virtual population with CHA_2_DS_2_-VASc score of 2. Sensitivity analyses were conducted by simulating limited OAC adherence (90%), permanent discontinuation after major bleeding and/or altered baseline bleeding risk in virtual RCTs.

### Statistical analyses

Five independent datasets were simulated with optimized parameters and mean ± standard deviation was calculated for all metrics. P values in K-M plots were derived by Log-rank test and hazard ratios (HRs) with 95% confidence intervals (95%CIs) were calculated in Cox regression models. Medians and 95%CIs of TTB and TTH analysis were estimated using Weibull models and 4000 Monte-Carlo simulations. Calibration bar plots with error bars, K-M plots, Cox regression models, and TTB/TTH analyses were conducted using ggplot2, survival, survminer, and brms packages in the statistical software R v4.3.0 (R core team, 2023) and R studio v2023.06.1.

## Results

### Patient-level AF model

A representative example of a virtual patient (**Figure 1B**) shows the model’s ability to capture all (a)symptomatic AF episodes during an individual’s lifetime, with significant AF progression from paroxysmal to persistent AF, and the occurrence of stroke, major bleeding and death events. Simulated continuous OAC was initiated after the virtual patient’s clinical AF diagnosis and either continued indefinitely or halted after the occurrence of a major bleeding (**Figure 1B, red boxes**).

### Model calibration and validation

We evaluated the extended model’s ability to reproduce key AF characteristics. The simulated AF incidence, prevalence, progression rate, episode duration and burden were consistent with clinical data (**Figure S2**). Importantly, when simulating study-specific patient inclusion criteria, the model accurately reproduced stroke incidence in the general population (**Figure 2A**), post-stroke acute fatality (**Figure 2B**), non-fatal (**Figure 2C**) and fatal major bleeding (**Figure 2D**) incidences in the general population without OAC, as well as major bleeding in AF patients without (**Figure 2E**) or with OAC (**Figure 2F**). Both stroke/TE and major bleeding incidence were significantly age-dependent in the general population.

**Fig. 2.**
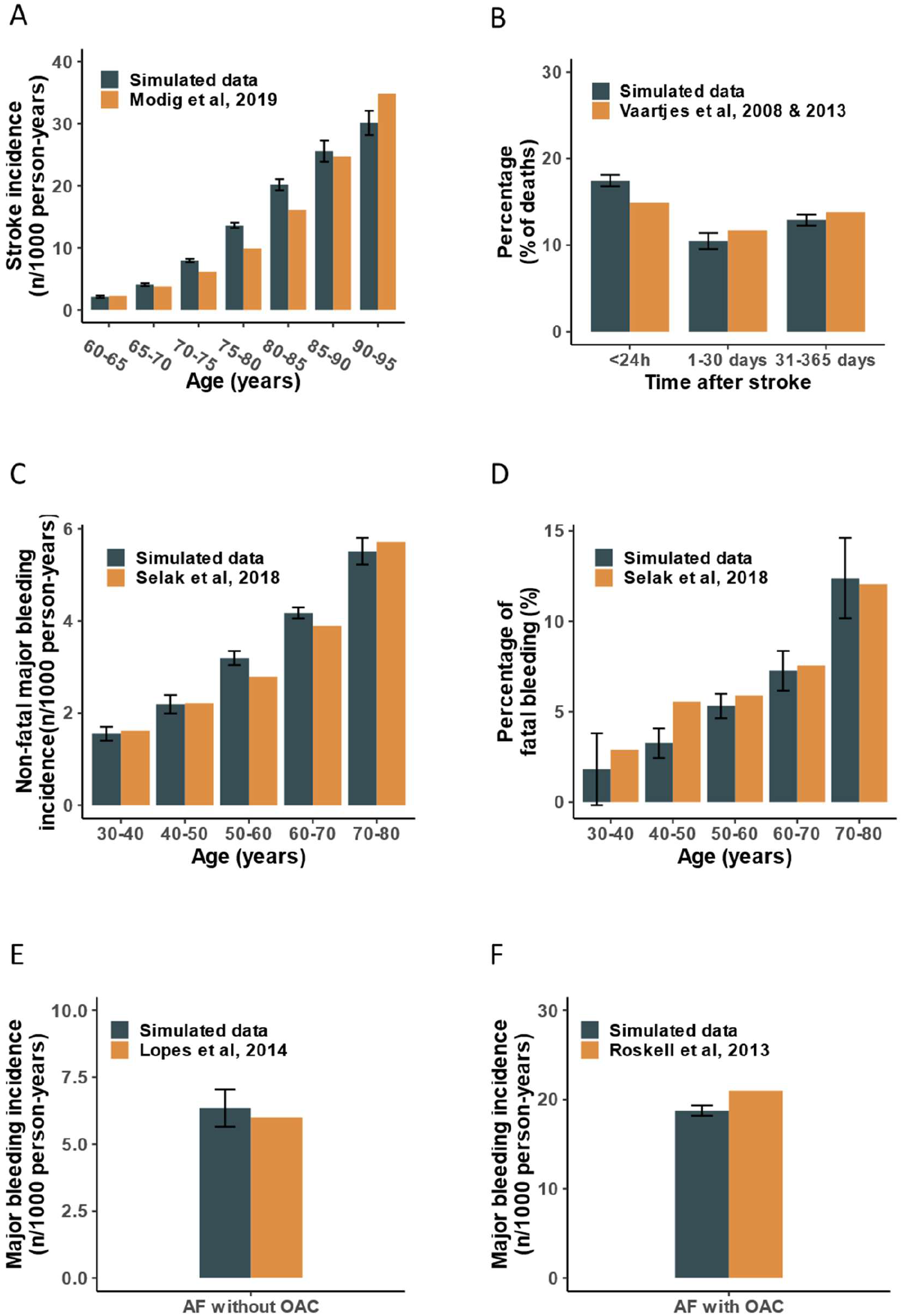
Study-specific **model calibration for stroke and major bleeding events. A**: Comparison between computed (black) and observed (orange) stroke incidence in the general population.^38^ **B**: Comparison between computed (black) and observed (orange) acute or long-term fatality after stroke in the general population.^40, 41^ **C-D**: The comparison between computed (black) and observed (orange) non-fatal major bleeding incidence (patient did not die within 28 days after major bleeding; **C**) and percentage of fatal major-bleeding in the general population without atrial fibrillation (AF) and oral anticoagulants (OAC; **D**).^39^ **E-F**: Comparison between computed (black) and observed (orange) major bleeding incidence in AF patients without OAC^42^ (**E**) or AF patients with OAC (**F**).^43^ All error bars are produced by 5 different repetitions with 10000 virtual individuals simulated.

Subsequently, stroke/TE incidence was further calibrated in virtual populations without (**Figure 3A**) and with OAC (**Figure 3B**), which showed excellent agreement between computed and observed incidences over a wide range of CHA_2_DS_2_-VASc scores. Independent model validation was conducted by comparing simulated and observed stroke/TE incidence in anticoagulation-naive AF patients with different CHA_2_DS_2_-VA scores from 0 to 5, reproducing the increasing trend of stroke/TE risk from 1 to 8 per 100 person-years (**Figure 3C**).

**Fig. 3.**
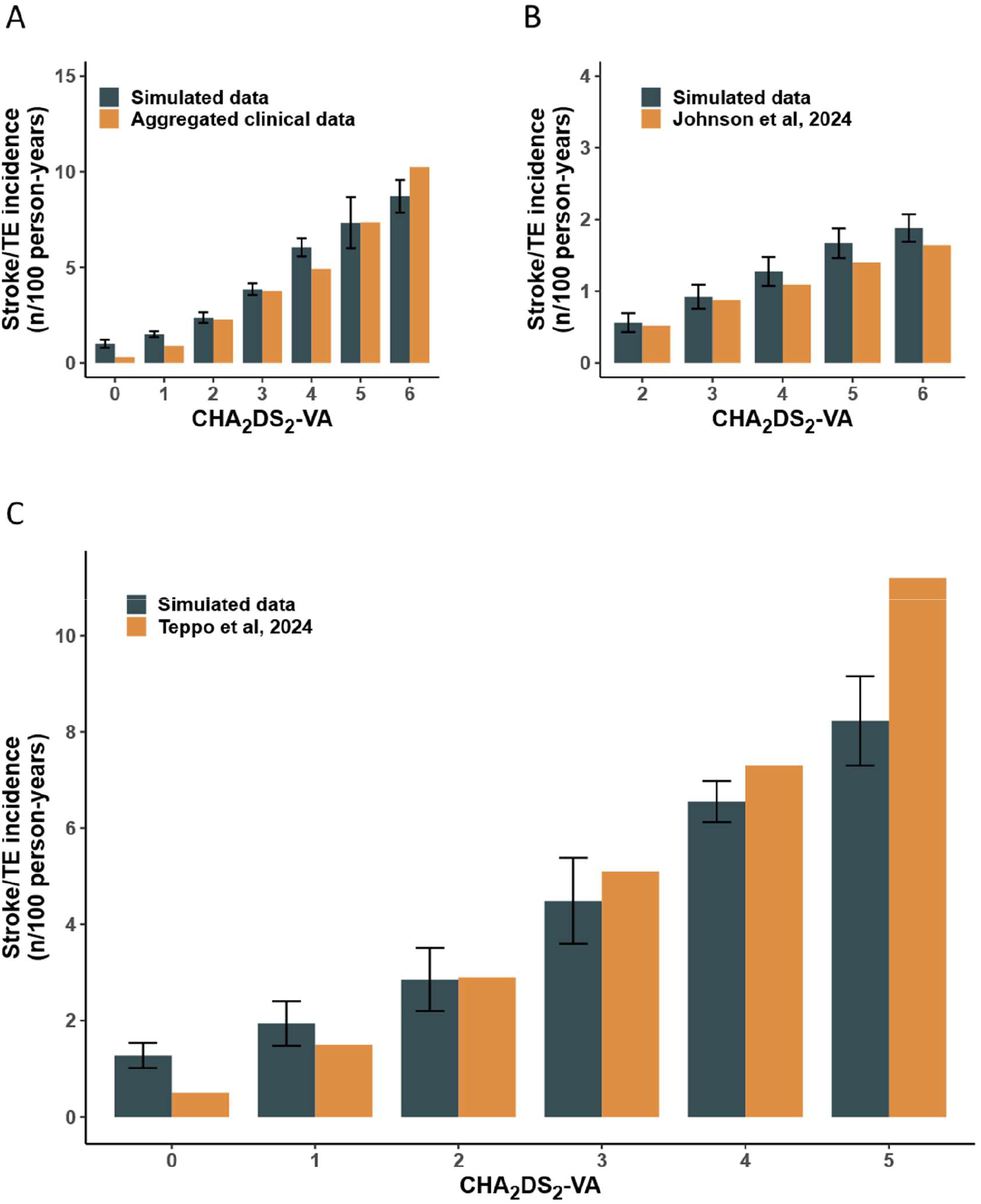
Stroke/thromboembolic events (**TE**) **in atrial fibrillation** (**AF**) **patients with different CHA**_2_**DS**_2_**-VASc scores and model validation using CHA**_2_**DS**_2_**-VA scores**. **A-B**: Comparison between computed and observed stroke/TE incidence in AF patients without^44–51^ (**A**) or with^52^ (**B**) oral anticoagulation across different CHA_2_DS_2_-VASc groups. **C**: Independent model validation of stroke/TE incidence for different CHA_2_DS_2_-VA scores in non-anticoagulated AF patients by simulating the inclusion criteria in Teppo et al, 2024.^53^ All error bars are produced by 5 different repetitions with 10000 virtual individuals simulated

### Benefits and harms of AF screening-guided OAC

The model was used to assess the benefits and harms of different AF screening strategies and subsequent OAC. In a virtual population with life-time follow-up and an average CHA_2_DS_2_-VA score of 2, K-M curves showed slightly lower cumulative stroke risk (P=0.002, **Figure 4A**) while significantly higher major bleeding risk in screening groups (P=0.001, **Figure 4B**), especially with LT-CRM (HR_LT-CRM_ (95%CI): 0.92 (0.88-0.96), P<0.001 for stroke; HR_LT-CRM_: 1.17 (1.07-1.27), P<0.001 for major bleeding compared to no screening). Sensitivity analyses showed that LT-CRM with screening-triggered OAC was associated with both significant stroke reduction and major bleeding increment across CHA_2_DS_2_-VA scores during a 35-year follow-up period (**Table S6**). However, these effects were diluted when simulating real-world OAC adherence, screening sensitivity, and permanent OAC discontinuation after bleeding (**Figure S3**, **Table S7**).

**Fig. 4.**
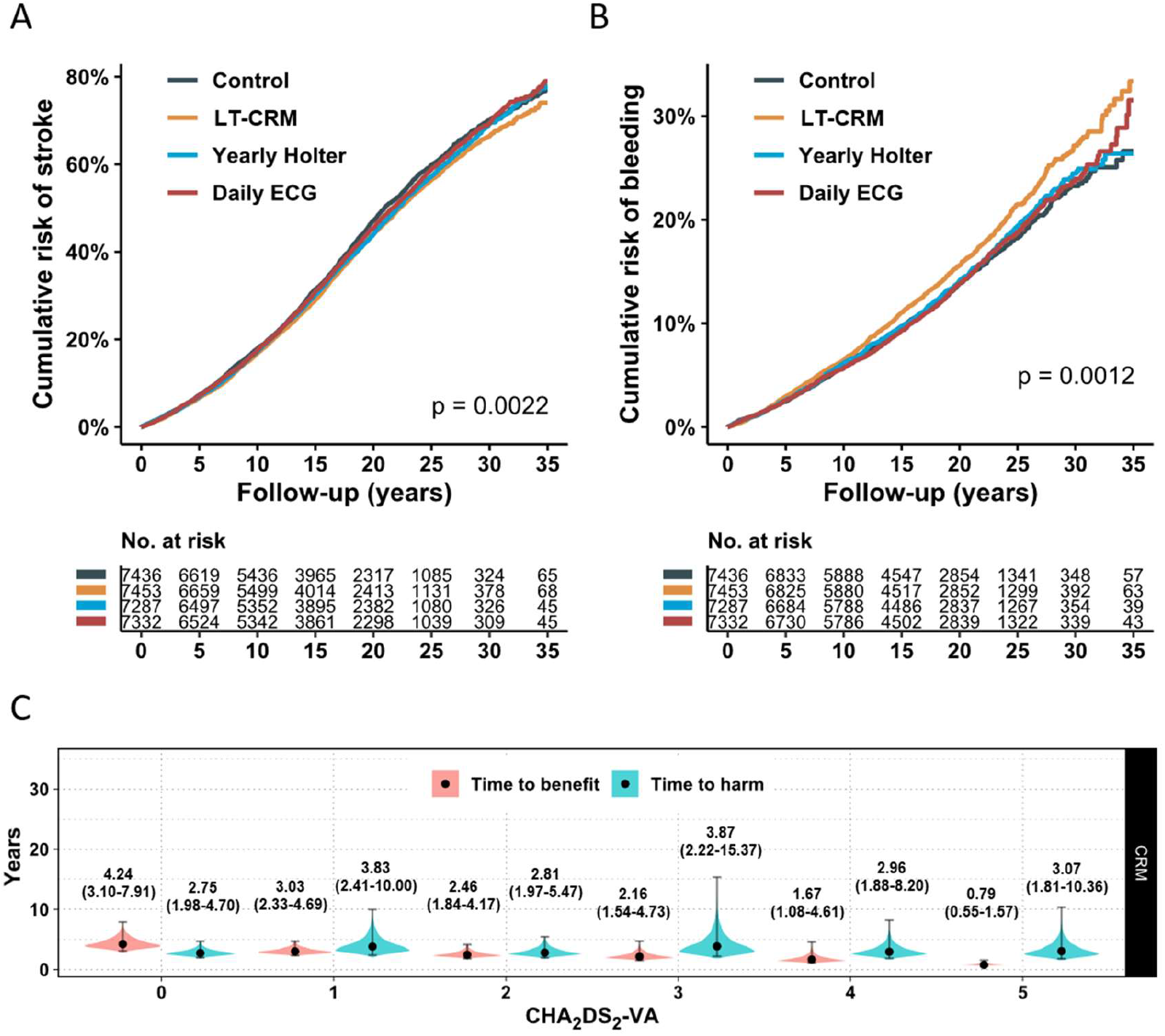
Risk-benefit assessment of three population-level systematic screening strategies with continuous oral anticoagulation therapy after diagnosed atrial fibrillation. A-B: Simulated cumulative risk of stroke (A) and major bleeding (B) in the control group without screening and systematic screening groups with a CHA_2_DS_2_-VA score of 2. P values in (A) and (B) are produced by Wald test of Cox regression models comparing screening groups to control groups with 10,000 individuals simulated in each arm. The numbers in risk tables are number of patients after selection based on the AF screening setting. C: Violin plots showing medians and 95% confidence intervals of time-to-benefit (stroke prevention) and time-to-harm (major bleeding) of LT-CRM with oral anticoagulation therapy at a threshold of 0.002 (i.e., 1 event in 500 screened individuals) across different CHA_2_DS_2_-VA scores. LT-CRM: continuous rhythm monitoring; Yearly Holter: yearly 14-day Holter monitoring; Daily ECG: daily three times 1-minute ECG monitoring.

TTB/TTH analyses with LT-CRM showed a progressive decrease in years needed to observe 1 fewer stroke event per 500 screened individuals with increasing CHA_2_DS_2_-VA scores, compared with similar years to observe 1 more major bleeding event. Consequently, TTB was markedly shorter than TTH in individuals with CHA_2_DS_2_-VA≥4 and significantly longer in individuals with CHA_2_DS_2_-VA=0 (**Figure 4C**). Real-world OAC adherence, screening sensitivity, and permanent discontinuation after bleeding did not change this pattern (**Figure S3**), suggesting a net benefit for screening in individuals with CHA_2_DS_2_-VA≥4.

### Continuous versus pill-in-pocket OAC strategies

The model was used to simulate the REACT-AF protocol and predict its outcomes, while adding an additional reference group without any OAC for comparison. The predicted occurrence of the composite primary endpoint (stroke or all-cause mortality) at 5 years in patients was significantly higher in non-anticoagulated patients with AF and CHA_2_DS_2_-VASc=2 compared with those with either pill-in-pocket or continuous OAC (log-rank P<0.001, **Figure 5A**), while no significant difference was identified between pill-in-pocket and continuous OAC (HR: 1.04, 95%CI: 0.93-1.16, P=0.48, **Table S8**). The risk of major bleeding at 5 years was significantly lower in AF patients without OAC compared to continuous OAC (HR: 0.41, 95%CI: 0.31-0.55, P<0.001; **Table S8;** log-rank P<0.001, **Figure 5B**). In the baseline model, pill-in-pocket OAC did not lower major-bleeding incidence compared to continuous OAC (HR: 0.92, 95%CI: 0.74-1.15, P=0.47; **Table S8**). Of note, when assessing the individual components of the primary endpoint, the stroke risk was also significantly higher (HR: 1.50, 95%CI: 1.17-1.92, P=0.0014) while mortality was similar (HR:0.98, 95%CI: 0.87-1.10, P=0.69) with pill-in-pocket versus continuous OAC (**Figure S4A-B** and **Table S8**). However, in virtual populations with doubled baseline bleeding risk, the difference in stroke incidence between continuous and rhythm-guided OAC was diminished (HR: 0.92, 95% CI: 0.71-1.19, P=0.52) and major bleeding was significantly lower (HR: 0.81, 95% CI: 0.68-0.96, P=0.014) in pill-in-pocket strategy compared with continuous OAC (**Figure S4C-D, Figure 5C-D** and **Table S9**). Sensitivity analyses considering perfect OAC adherence and no OAC discontinuation after major bleeding showed higher stroke (HR: 1.84, 95% CI: 1.38-2.45, P<0.001) and lower major bleeding (HR: 0.80, 95% CI: 0.65-0.99, P=0.04) in pill-in-pocket compared with continuous OAC (**Figure S5-6** and **Table S10**). Importantly, lower OAC adherence significantly reduced the differences in stroke and major bleeding risks between both strategies, but the influence on stroke was larger than that on major bleeding (**Figure S7**). These simulations of an ongoing RCT suggest that rhythm-guided OAC might be beneficial in certain populations.

**Fig. 5.**
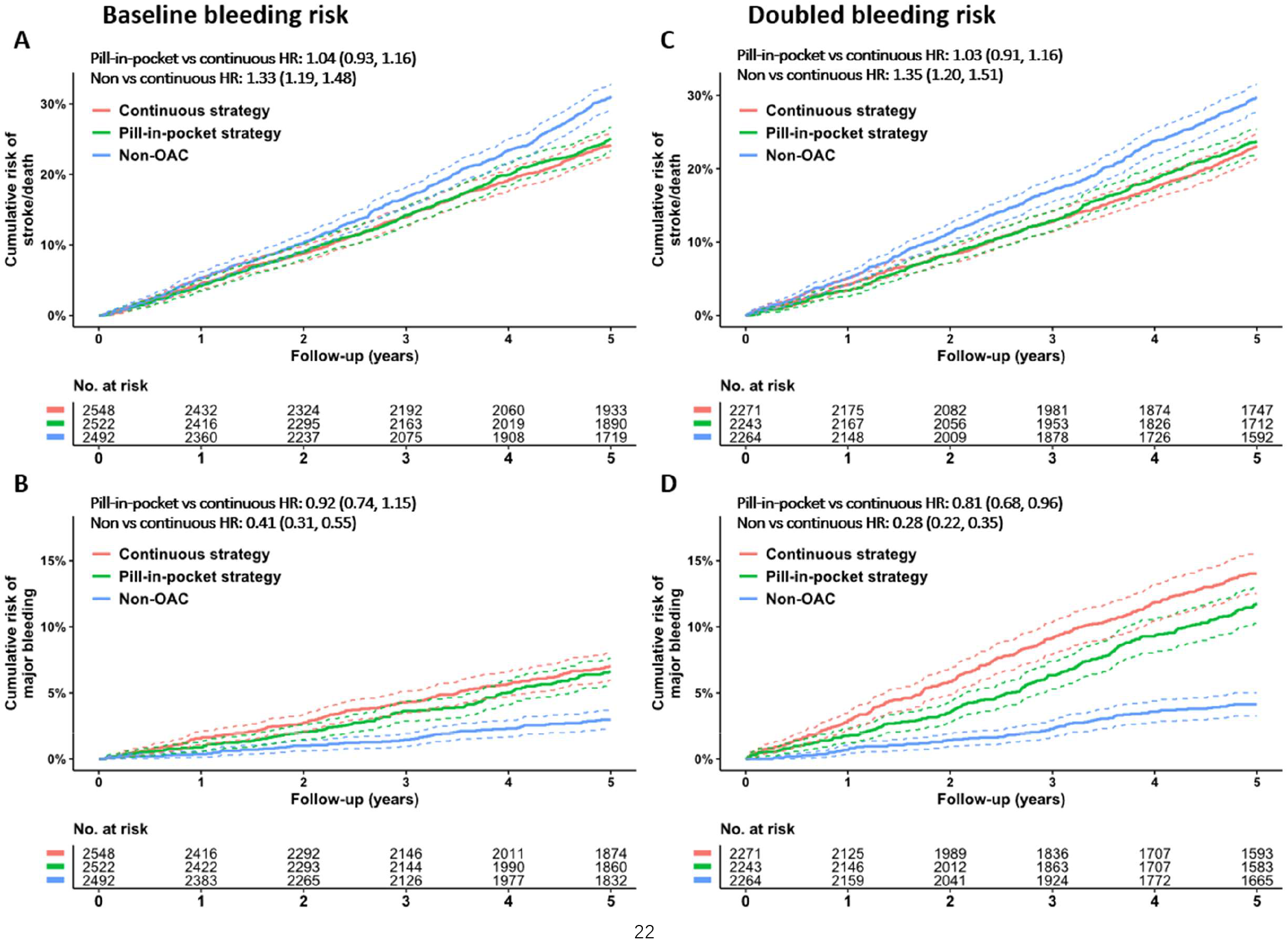
Predicted stroke/mortality and major bleeding risk of different oral anticoagulation (OAC) strategies based on the REACT-AF trial, with 90% OAC adherence and OAC discontinuation after major bleeding. A-B: Comparison of predicted stroke/mortality risk (A) and bleeding risk (B) in patients without oral anticoagulation, standard continuous oral anticoagulation or a pill-in-pocket oral anticoagulation strategy with rhythm-guided time-delimited OAC defined by the REACT-AF trial.^21^ C-D: The same comparison in virtual populations with doubled baseline bleeding risks. All P values are produced by Wald test of Cox regression models comparing pill-in-pocket or none OAC strategy to continuous OAC strategy with 10000 individuals simulated at beginning in each arm. The numbers in risk tables are number of patients after selection against inclusion criteria of REACT-AF trial.

## Discussion

We presented a well-calibrated computational model incorporating lifetime AF episodes, disease progression, and patient-level clinical outcomes.^17^ Virtual screening RCTs employing this model suggested that LT-CRM-initiated OAC led to modest long-term stroke reduction and significantly increased major bleeding across CHA_2_DS_2_-VA scores from 0 to 5. These effects were influenced by real-world limitations, including imperfect OAC adherence, screening sensitivity, and OAC discontinuation after major bleeding. In populations with CHA_2_DS_2_-VA≥4, TTB was much shorter than TTH, indicating a net benefit of LT-CRM-initiated OAC that was not present with lower CHA_2_DS_2_-VA. Furthermore, a rhythm-guided pill-in-pocket OAC strategy was non-inferior to standard continuous OAC for the combined endpoint of stroke and mortality in patients with CHA_2_DS_2_-VASc=2, but was superior in major bleeding prevention in patients with high bleeding risk.

### Strengths of the current model

While computational models have been commonly used to simulate the outcomes of OAC therapy, e.g., to perform cost-effectiveness analyses,^22, 23^ these approaches lacked dynamic modeling of individual AF episodes, limiting their suitability for evaluating AF screening. Other modeling approaches used to analyze benefits of AF screening,^24, 25^ similarly omitted individual AF episodes or AF progression, missing key elements of AF pathophysiology and precluding the assessment of rhythm-guided OAC therapy. Our model is the first to capture the dynamic and progressive occurrence of AF episodes and associated outcomes with or without OAC therapy, enabling simulation of virtual RCTs comparing different OAC therapies.

The stroke-reduction benefit of systematic AF screening remains uncertain, while concerns persist regarding the potential major bleeding risk after screening-triggered OAC.^4, 6, 7, 9, 26^ Although pooled data showed a modest 7% reduction in stroke/systemic embolism without significant impacts on major bleeding or all-cause mortality, larger RCTs with longer follow-up would be needed to confirm these results and identify the subgroups with the largest benefit.^27^ However, such RCTs are costly and time-consuming, hindering advances in clinical practice. Our well-calibrated and independently validated model provides an efficient and cost-free method for risk/benefit analyses of AF screening and OAC therapy, enabling long-term follow up of virtual patients and head-to-head comparison of therapeutic strategies.

### Clinical implications: role of screening-induced OAC in AF

Our simulations revealed modest long-term stroke reduction with LT-CRM-initiated OAC across CHA_2_DS_2_-VA scores. Other screening modalities including daily 1-minute ECGs and yearly 14-day Holter monitoring had minimal impact on stroke prevention, consistent with the relatively low yield of AF screening in our previous simulations^17^ and clinical data.^7, 9^ Although simulated LT-CRM-based screening decreased stroke risk, it also strongly promoted major OAC-related bleeding. This is consistent with findings from RCTs in subclinical AF patients with continuous monitoring and OAC,^11, 12, 19^ and supports the conclusion from the LOOP study that not everyone benefits from screening and not every screening-detected AF benefits from OAC.^6^ Better risk stratification is urgently needed to identify subgroups who may benefit from continuous screening. Baseline stroke risk (CHA_2_DS_2_-VA scores) may help to improve the risk/benefit profiles of such interventions. Our analyses showed a progressive shortening of TTB and comparable TTH of LT-CRM-initiated OAC with increasing CHA_2_DS_2_-VA scores, leading to significantly shorter TTB than TTH in those with CHA_2_DS_2_-VA≥4. This finding is consistent with the conclusion from a subgroup analysis of ARTESIA, showing that continuous OAC might be beneficial in patients with CHA_2_DS_2_-VASc≥4 and subclinical AF detected by continuous monitoring devices.^28^ It should be noted that these analyses assume perfect OAC therapy. In the analyses accounting for real-world OAC adherence, screening sensitivity, and OAC discontinuation after major bleeding, the between-group difference of stroke and major bleeding risks observed were diluted across CHA_2_DS_2_-VA scores for all screening strategies, although the general trend remained. Together, these findings argue for a risk-stratified approach that prioritizes individuals most likely to derive net benefit, e.g., based on CHA_2_DS_2_-VA scores. Moreover, our simulations show that the risk-benefit profiles of AF screening will strongly depend on the interaction of real-world OAC adherence, post-bleeding OAC discontinuation, and screening technology accuracy, which are difficult to control in real-world studies.

### Clinical implications: role of pill-in-pocket OAC in AF

Besides enabling screening for previously undiagnosed AF, wearable devices have enabled better monitoring of patients with known AF, potentially enabling rhythm-based, time-delimited (pill-in-pocket) OAC to reduce major bleeding risk. Several pilot studies with pill-in-pocket OAC have shown promising results,^29–35^ giving rise to the large REACT-AF trial. REACT-AF is an ongoing non-inferiority study comparing pill-in-pocket OAC with usual, continuous OAC in terms of stroke or all-cause mortality (primary endpoint) and major bleeding prevention (secondary, superiority endpoint). However, REACT-AF results will only be available in 2029.^36^ By simulating the REACT-AF protocol, our model predicts that a pill-in-pocket OAC strategy results in a significant 20% reduction in major bleeding while maintaining a similar incidence of the combined primary endpoint of stroke and all-cause mortality compared with the traditional continuous OAC after clinical AF diagnosis, at least in patients with a sufficiently large bleeding risk. Additionally, our model enabled a comparison to virtual patients without OAC, something that would be unethical in real-world studies, to provide insights into the absolute risk/benefit of each approach, showing a significantly higher risk of stroke and a lower risk of major bleeding compared with either OAC strategies.

These results suggest that REACT-AF might successfully achieve the predefined non-inferiority in efficacy and superiority in safety of pill-in-pocket strategy, indicating that the potential adverse effects of continuous OAC usage, especially in those with low AF burden and low stroke risks, can be partially ameliorated by rhythm-guided anticoagulation therapy. However, our sensitivity analyses show that the differences between intermittent OAC and continuous OAC depend on bleeding risk, OAC adherence and post-bleeding OAC discontinuation. Thus, real-world OAC treatment limitations may dilute or even overturn the effects derived from RCTs. These results highlight the importance of carefully monitoring long-term safety of this new OAC approach and a potential need to identify subgroups of patients with preferential risk/benefit profile. Discrimination based on overall AF burden might represent an interesting strategy based on recent data suggesting its modulating role on stroke risk.^37^ Moreover, the quantitative information provided by the model may facilitate shared decision making on the potential trade-off between increased stroke risk and decreased major bleeding risk.

Importantly, at present, our model predictions are fully hypothesis-generating and not direct evidence to support clinical implementation. While computational models are valuable for exploratory analysis in costly, infeasible, or unethical scenarios, and may support trial design by comparing different options, they cannot replace real-world RCTs. Our modeling results should be validated against the outcomes of real trials, including REACT-AF, when the results are available.

### Limitations

Some limitations of the current study have to be considered. Firstly, the components underlying the CHA_2_DS_2_-VASc / CHA_2_DS_2_-VA scores were not explicitly modeled. Instead, all different risk factors were integrated into a single parameter modifying the baseline stroke risk. As such, the current model can only capture the average population-level stroke risk dynamics rather than the individual risk profiles and the interpretation of these scores is somewhat different from the traditional risk evaluation for an individual patient with AF. Thus, results should be interpreted cautiously as population-level inferences. Secondly, major bleeding and stroke were considered as irreversible processes and recurrent events were ignored in the current model. However, only time to the first occurrence of clinical outcomes was used for data analysis in our study, in line with previous clinical trials, limiting the impact of this limitation.^38, 39^ Thirdly, the model focused only on OAC initiation after either clinical or screening-detected AF diagnosis, without considering risk-factor modification or rhythm control, which are fundamental components of comprehensive AF management.^1^ This limitation might underestimate the benefits of AF screening and pill-in-pocket OAC strategy on stroke and bleeding risk, although the simulated results reflect the situation evaluated in most recent trials.

### Conclusions

The model confirms that OAC treatment after systematic AF screening using LT-CRM is a double-edged sword with a higher major bleeding risk offsetting a lower stroke incidence. Individuals with CHA_2_DS_2_-VA≥4 may benefit from systematic AF screening followed by OAC initiation. A LT-CRM-guided pill-in-pocket OAC strategy after AF diagnosis may significantly reduce the major bleeding risk while maintaining a similar combined primary outcome of stroke and all-cause mortality after 5 years follow-up. Limitations of real-world OAC treatment and screening technology may interact with the risk-benefit profiles of screening-controlled OAC and may dilute the outcomes observed in RCTs. More generally, this study provides the first computational prediction of the outcome of an ongoing OAC RCT and shows how our approach can be used in the future to facilitate clinical trial design.

## Data Availability

The model and all original code will be deposited at https://github.com/HeijmanLab/ to allow others to use and expand this novel approach.

## Acknowledgements

The authors thank Chuanyi Huang and Xin Zhou for insightful statistical suggestions, and Paul Volders for helpful discussions.

